# Objective Predictors of Visual Quality of Life in Parkinson’s Disease

**DOI:** 10.64898/2026.03.25.26349331

**Authors:** Rhea Mehta, Pooja Nambiar, Camilla Kilbane, Fatema F Ghasia, Aasef G. Shaikh

**Author notes:** **Corresponding Author:** Aasef G. Shaikh,MD,PhD Department of Neurology University Hospitals, 11100 Euclid Avenue Cleveland, OH 44110.

## Abstract

**Background:** Visual dysfunction is a common but underrecognized contributor to disability in Parkinson’s disease (PD), particularly deficits in binocular vision and vergence that impair reading, near work, and quality of life. The relationship between objective oculomotor abnormalities and patient⍰reported visual disability remains incompletely understood.

**Methods:** We studied 25 individuals with PD and 11 age⍰matched controls who completed the National Eye Institute Visual Function Questionnaire⍰25 (VFQ⍰25) and the Convergence Insufficiency Symptom Survey (CISS). Participants underwent comprehensive clinical ophthalmologic assessment and high⍰resolution binocular eye tracking to quantify vergence latency, gain, fixation dynamics, and drift variability. Associations between objective measures and patient⍰reported outcomes were examined, and predictive models were developed using clinic⍰only and combined clinic + eye⍰tracking approaches.

**Results:** Compared with controls, PD participants demonstrated significantly worse VFQ⍰25 composite scores and higher CISS scores, driven primarily by impairments in near activities and mental health. Clinically, PD was characterized by convergence insufficiency rather than generalized visual loss. Objective eye tracking revealed delayed vergence initiation, reduced gain, and increased instability. In PD, both clinical convergence measures (notably near⍰point convergence) and dynamic eye⍰tracking metrics strongly correlated with VFQ⍰25 and CISS scores, whereas such relationships were absent in controls. Predictive models showed limited performance using clinic measures alone, but improved with inclusion of eye⍰tracking variables.

**Conclusions:** Visual disability in PD is tightly linked to convergence insufficiency and dynamic oculomotor instability. Simple clinical measures such as near⍰point convergence, augmented by eye tracking when available, provide meaningful insight into patient⍰reported visual quality of life.

## Introduction

Parkinson’s disease (PD) is a progressive neurodegenerative disorder affecting approximately 10 million people worldwide. Although traditionally characterized by its motor manifestations, including tremor, bradykinesia, and rigidity, PD is increasingly recognized as a multisystem disorder with prominent non⍰motor features. Among these, visual dysfuntion represents a common yet underappreciated source of disability. Visual abnormalities in PD include deficits in eye movement control such as vergence (convergence when shifting gaze from far to near and divergence when shifting from near to far), rapid gaze shifts (saccades), and gaze stabilization mechanisms, including saccadic intrusions^1–7^.

Vergence eye movements are particularly relevant to quality of life in PD. Reading difficulty is among the most frequent visual complaints reported by patients, and objective measures of reading performance are consistently poorer in individuals with PD compared with age⍰matched controls. Convergence insufficiency may contribute to diplopia, particularly during near work such as reading^8–10^. Efficient visual scanning during real⍰world tasks such as reading requires both accurate saccadic eye movements and continuous vergence adjustments to maintain binocular alignment across varying viewing distances. Diplopia itself is reported in up to 30% of patients across studies and is associated with disease severity, motor fluctuations, cognitive and neuropsychiatric features, and treatment state^10,11^. Vergence also plays a critical role in depth perception and postural stability. Falls occur at a higher frequency in PD, and vergence insufficiency may represent a significant contributor to fall risk beyond motor impairment alone^12^. Many PD patients have previously unrecognized ocular misalignment or convergence insufficiency on formal examination, and a subset may benefit from targeted interventions such as optimized dopaminergic therapy, optical correction, or orthoptic treatment^8,9,13,14^. These findings underscore that vergence insufficiency and diplopia should not be regarded as minor secondary complaints; rather, they constitute a substantial component of the visual burden of PD, interfering with reading, mobility, and daily functioning, and contributing to impaired vision⍰related quality of life documented across clinical, questionnaire⍰based, and ophthalmologic studies^8,9,15–18^.

Given the prevalence and impact of visual impairments in PD, patient⍰reported outcome measures offer a critical window into the lived visual experience of this population. The National Eye Institute Visual Function Questionnaire⍰25 (VFQ⍰25) is widely used to assess the relationship between visual function and quality of life across twelve subscales, including near and distance activities, social functioning, mental health, dependency, and peripheral vision^19^. Studies consistently demonstrate that VFQ⍰25 composite scores are significantly reduced in individuals with PD compared with controls^8,9,15^, with both studies also reporting a high prevalence of convergence insufficiency. Furthermore, a greater number of significant correlations have been observed between clinical measures of visual function—such as low⍰ and high⍰contrast visual acuity—and VFQ⍰25 subscales in PD compared with control populations^17^.

Undetected ophthalmologic disorders, such as ocular misalignment and convergence insufficiency, are linked to worse VFQ⍰25 scores and increased fall risk in PD^9^. Higher⍰order visual processes, including perception, visual construction, attention, and processing speed, exert an even stronger influence on perceived visual disability and quality of life than basic sensory deficits^17^. Reading difficulty, impaired focusing, glare sensitivity, diplopia, and slowed visual processing consistently emerge as key expressions of reduced visual quality of life in PD^14,16,20^. Collectively, this literature demonstrates that visual disability in PD is common, multidimensional, clinically meaningful, and insufficiently captured by routine neurologic or ophthalmologic assessments alone.

Despite substantial prior work, objective visual abnormalities and patient⍰reported visual quality of life in PD are still typically examined in parallel rather than in direct relation to one another. Objective clinical and oculographic measures quantify physiologic deficits in vergence and binocular alignment, whereas instruments such as the VFQ⍰25 and CISS capture the lived burden of visual dysfunction. Without integrating these domains, the functional significance of specific examination findings—and their contribution to patients’ visual disability—remains unclear. Establishing these relationships is essential for distinguishing abnormalities that are merely detectable from those that meaningfully affect daily functioning. Moreover, objective, standardized clinical and eye⍰tracking measures offer sensitivity to subclinical dysfunction and may serve as reproducible biomarkers of quality of life, particularly if they can predict patient⍰reported outcomes. Such predictive links would have direct translational value by enabling risk stratification, guiding targeted interventions, monitoring therapeutic response, and focusing treatment on deficits most likely to improve patient⍰centered outcomes.

Accordingly, this study aims to (1) characterize the impact of visual dysfunction on everyday activities using the VFQ⍰25 and CISS; (2) comprehensively assess objective ocular motor capacity in PD through clinical and oculographic measures of binocular alignment and vergence control; (3) determine which objective parameters correlate with vision⍰related quality of life; and (4) develop predictive models to identify the measures that best forecast patient⍰reported visual outcomes. Identifying such objective physiomarkers will inform therapeutic strategies in PD and ultimately support efforts to improve patients’ quality of life.

## Methods

We studied 36 participants, including 25 with PD defined by UK Brain Bank Clinical Criteria and 11 healthy controls. The mean age of PD was 69±8 years. All patients were consented to the standards of the Declaration of Helsinki. The protocol was approved by Cleveland Clinic Institutional Review Board, the participants signed on approved informed consent form. During each individual visit, the participants completed both the VFQ-25 and CISS, either in the interview-administrated format (interviewer to read off the questions through a standardized script) or independently using the self-administered format. Both formats were identical in their content. The results of the VFQ-25 were interpreted with well-established conversion protocol, resulting in numerical scores out of 100 for the aforementioned 12 sub-scale categories and a total composite score^19^. For all these thirteen scores, a score of 100 meant no negative impact on the quality of life. CISS questionnaires were also converted to total numerical scores based on the Likert scale included within the survey, with scores from 0 - 4 representing symptom frequency from “never” to “always.” Higher total CISS score signified worsened symptom severity^21^.

All participants had objective eye movement assessments on the same day as answering the questionnaire CISS and VFQ-25. The participants were seated in a chair with chin and forehead secured in front of the ocular tracking system independently measuring both eyes movements in horizontal and vertical directions (500 Hz sampling g frequency and 0.1 degree spatial resolution; EyeLink 1000^TM^, SR Research, Mississauga, ON); see our lab’s previous publications for details ^22–25^. To summarize, each session began with three-point calibration in the following horizontal and vertical components subtended by the binocular eye positions: 0°and 10°, 15° and 10°, 15° and 10°. In order to assess vergence, the light emitting diode (LED) targets were placed within a sagittal plane at 20, 55, 150, and 244 cm. Convergence was measured as the participants moved their gaze to a near LED target from one farther away and then vice versa for divergence. Vergence component latency was recorded as an eye movement shift from baseline at least 0.5° with a velocity greater than 3°/s following shift in target. Vergence gain was captured as a ratio of the actual change in the vergence amplitude and desired change in the vergence amplitude (i.e., the vergence demand).

Patient⍰reported outcomes included the VFQ⍰25 composite score and the CISS. Ocular⍰motor predictors were mean convergence/divergence component latencies, mean convergence/divergence total gaze⍰shift gains, and near-point convergence (NPC). Visual acuity was converted from Snellen notation to logMAR and averaged across eyes. Stereopsis values were extracted from free⍰text entries, with the worst value retained and log⍰transformed. Covariates included age and gender.

Descriptive statistics were computed by group. PD–control comparisons used Welch’s t⍰tests with Cohen’s d. Multivariable analyses employed standardized ordinary least squares regression with bootstrap confidence intervals (3,000 resamples).

Multicollinearity was assessed using variance inflation factors, guiding model reduction. Subset analyses used k⍰fold cross⍰validation (K = 5 in PD; leave⍰one⍰out in controls) to evaluate generalizability. Pearson correlations with Fisher z confidence intervals were used to relate clinical measures to outcomes within PD.

## RESULTS

### Cohort Characteristics and Data Overview

A total of 36 participants were included in this study, comprising 25 PD patients and 11 healthy controls. The dataset encompassed a multidimensional characterization of visual function, including:

1) Disease-specific measures in PD such as Unified Parkinson’s Disease Rating Scale part 3 (UPDRS-3), disease duration, and daily levodopa dose
2) Patient-reported outcomes, including the VFQ-25 composite and subscales as well as the CISS.
3) Clinically assessed visual function, including near point convergence (NPC) across multiple methods, stereopsis, ocular alignment (near and distance exodeviation), and other standard ophthalmologic measures.
4) High-resolution eye-tracking metrics, capturing dynamic vergence control through measures of latency, gain, drift variability, fixation stability, and vergence strategy utilization.

Within the PD cohort, disease severity (UPDRS-3), levodopa dose, and age were not significantly associated with visual quality of life or symptom burden, indicating that visual dysfunction represents a partially independent domain of disease expression. Disease duration demonstrated a modest, non-significant trend toward association with VFQ-25 scores (ρ = −0.35, p = 0.086).

### Group Differences in Visual Quality of Life and Symptom Burden

Comparative analysis between PD patients and healthy controls revealed significant impairments in vision-related quality of life and increased symptom burden in PD. Specifically, VFQ-25 composite scores were significantly lower in PD (86.54 ± 12.14) compared to controls (93.97 ± 5.78; t = −2.49, p = 0.018, FDR-adjusted p = 0.043) (**Table 1**), corresponding to a moderate-to-large effect size (Cohen’s d = −0.70). In contrast, CISS scores were markedly higher in PD (16.88 ± 10.02 vs 4.82 ± 6.91; t = 4.17, FDR-adjusted p = 0.002), with a large effect size (d = 1.31), indicating substantially increased binocular visual discomfort (**Table 1**).

**Table 1.**
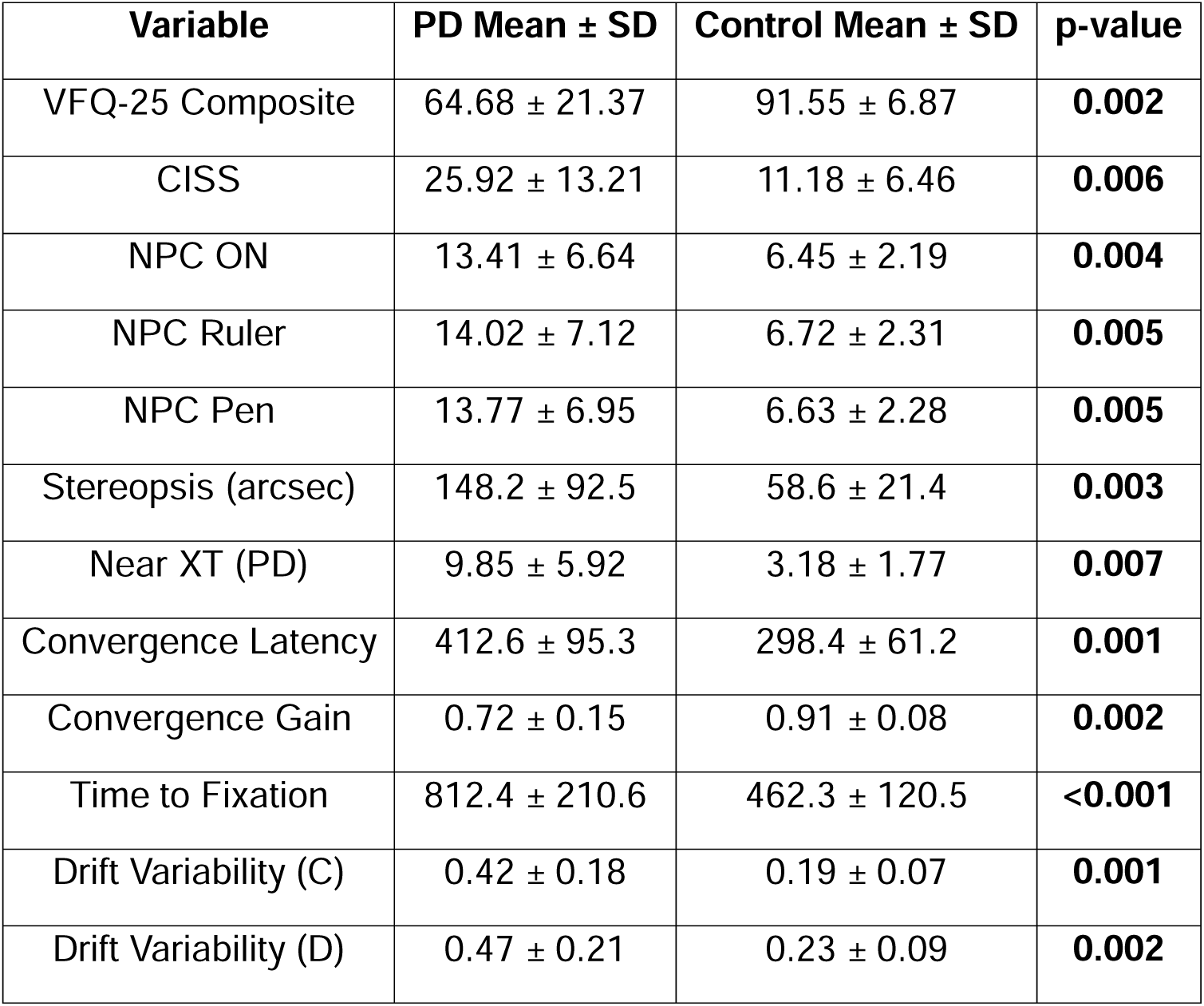

Subdomain analysis of the VFQ-25 demonstrated that these impairments were not uniform but concentrated in specific functional and psychosocial domains. PD patients had significantly lower scores in Near Activities (82.83 ± 19.41 vs 96.97 ± 7.70; p = 0.004, FDR p = 0.015, d = −0.84), Mental Health (81.75 ± 23.38 vs 96.88 ± 6.07; p = 0.005, FDR p = 0.016, d = −0.75), and General Health (42.00 ± 22.50 vs 77.27 ± 20.78; FDR p = 0.002, d = −1.60), with additional moderate effects observed in Role Difficulties (d = −0.58) and Social Functioning (d = −0.49) (**Figure 1**). In contrast, domains such as General Vision, Dependency, and Driving did not differ significantly between groups after correction for multiple comparisons, indicating relative preservation of these aspects of visual function in this sample (**Figure 1**).

**Figure 1.**
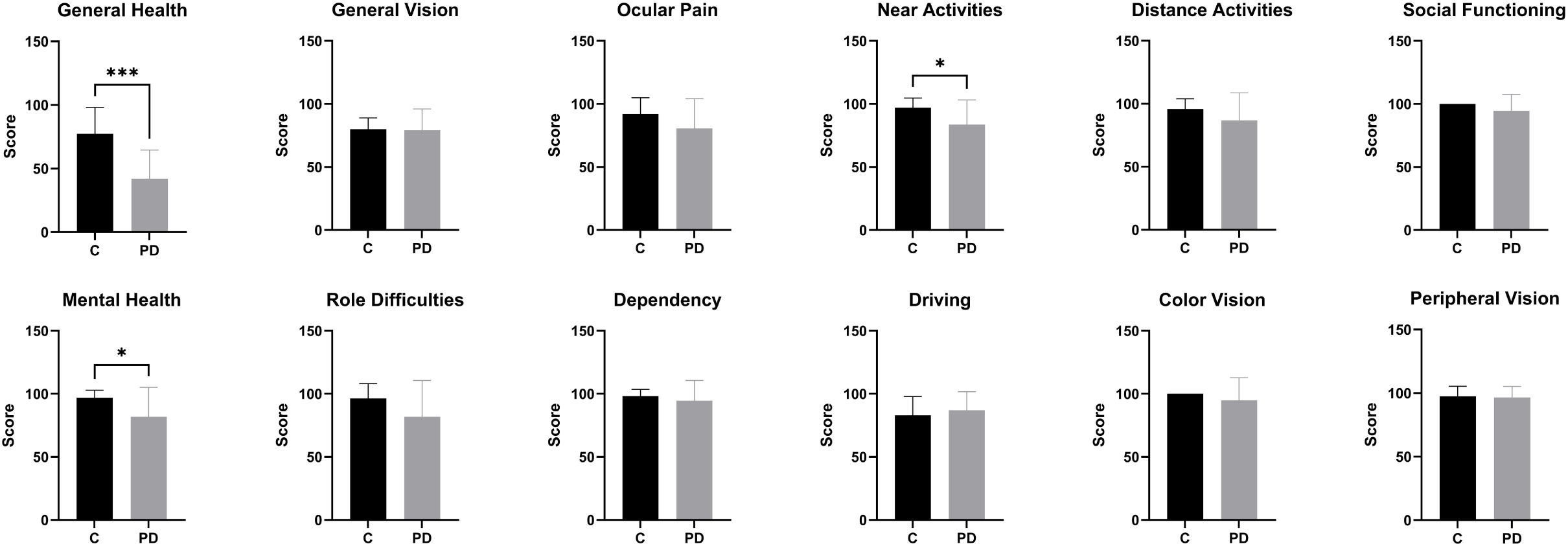
Mean subscale scores for control and PD groups. * signifies statistically significant difference and quantity represents relative value of p value.

### Clinical Measures of Visual Function

Clinical assessments demonstrated that the primary visual deficit in PD was convergence insufficiency rather than generalized visual impairment. NPC was consistently increased in PD across measurement methods, including NPC (17.40 ± 9.40 vs 11.40 ± 5.68; p = 0.029, d = 0.70), NPC Ruler (d = 0.74), and NPC Pen (d = 0.77), indicating a moderate-to-large impairment in convergence ability (**Table 1**). Additional measures of near visual function, including RAF Ruler (19.00 ± 7.48 vs 10.50 ± 5.01; p = 0.010, d = 1.24) and Distance Pen (16.67 ± 7.23 vs 8.58 ± 5.19; p = 0.013, d = 1.20), demonstrated large effect sizes, further supporting substantial deficits in near visual performance (**Table 1**).

In contrast, measures of basic visual function, including color vision and peripheral vision, did not differ significantly between groups (all FDR-adjusted p > 0.20), and stereopsis was not significantly different, likely reflecting variability and limited sample size in this measure. Age was also comparable between groups (69.48 ± 7.78 vs 69.55 ± 6.56; p = 0.979), indicating that observed differences were not attributable to demographic confounding.

### Eye-Tracking Measures of Vergence Control

Eye-tracking analyses revealed robust and consistent abnormalities in dynamic vergence control in PD. Patients demonstrated significantly prolonged convergence component latency (0.815 ± 0.427 vs 0.541 ± 0.193; p = 0.017, FDR p = 0.047, d = 0.72) and divergence component latency (1.065 ± 0.706 vs 0.583 ± 0.122; p = 0.003, FDR p = 0.012, d = 0.79), indicating delayed initiation of vergence movements (**Table 1**).

In parallel, convergence gain was significantly reduced in PD (0.568 ± 0.425 vs 0.855 ± 0.180; p = 0.010, FDR p = 0.030, d = −0.76), as was divergence gain (−0.445 ± 0.416 vs −0.811 ± 0.277; p = 0.008, FDR p = 0.027, d = 0.95), reflecting impaired accuracy of eye alignment. Gain variability, measured with standard deviation, was also increased in PD (0.302 ± 0.279 vs 0.135 ± 0.133; p = 0.026, FDR p = 0.065, d = 0.67), suggesting instability in vergence execution (**Table 1**).

Collectively, these findings demonstrate that PD is characterized by both slowed vergence initiation and reduced movement precision, consistent with impaired dynamic oculomotor control.

### Associations Between Visual Quality of Life and Clinically Measured Vergence and Alignment

Across the PD cohort, subjective visual quality of life was tightly linked to objective deficits in binocular vision, particularly convergence insufficiency. The VFQ-25 Composite score showed moderate correlations with multiple clinical markers of near binocular dysfunction. Specifically, worse VFQ-25 scores were associated with larger NPC distances (r ranging from approximately −0.45 to −0.55), indicating that individuals who required a greater distance to achieve convergence reported poorer visual functioning. Similar associations were observed for NPC Pen (r ≈ −0.40 to −0.50) and NPC Ruler, reinforcing the robustness of this relationship across measurement modalities (**Table 2**).

**Table 2.**

| **VFQ-25** |  |  |
| --- | --- | --- |
| **Variable** | **$\rho$** | **p-value** |
| Time to Fixation | −0.68 | **<0.001** |
| Drift Variability | −0.64 | **<0.001** |
| Convergence Gain | 0.62 | **<0.001** |
| Convergence Latency | −0.55 | **0.002** |
| NPC ON | −0.42 | **0.03** |
| Stereopsis | −0.38 | **0.04** |
| Disease Duration | −0.35 | **0.086** |
| **CISS** |  |  |
| **Variable** | **$\rho$** | **p-value** |
| Time to Fixation | 0.66 | **<0.001** |
| Drift Variability | 0.61 | **<0.001** |
| Latency Variability | 0.52 | **0.004** |
| Near XT | 0.39 | **0.04** |

Measures of ocular alignment also contributed meaningfully. Near exodeviation (Near XT) correlated negatively with VFQ 25 Composite (r ≈ −0.35 to −0.45), suggesting that greater outward deviation at near was linked to worse perceived visual ability (**Table 2**). Distance Pen and RAF Ruler—both reflecting near point of accommodation and convergence—also showed moderate negative associations (r ≈ −0.40 to −0.45) (**Table 2**).

Subscale analyses revealed a specific vulnerability of near vision–dependent activities. The Near Activities subscale demonstrated the strongest clinical coupling, with correlations of approximately −0.55 with NPC and −0.50 with Near XT. These values indicate that patients with the most pronounced convergence insufficiency experienced the greatest functional impairment in reading, close work, and other near vision tasks.

Symptom burden, quantified by the CISS, showed even stronger associations with clinical convergence measures. CISS correlated positively with NPC (r ≈ 0.55–0.65) and Near XT (r ≈ 0.45–0.55), indicating that patients with poorer convergence and greater near exodeviation reported substantially more symptoms of eye strain, diplopia, and near vision fatigue.

Taking together, these findings demonstrate that clinical markers of convergence insufficiency are the primary drivers of subjective visual disability in PD, with consistent effects across both functional (VFQ-25) and symptom based (CISS) measures. In contrast, controls exhibited minimal structure in clinical–subjective associations. VFQ-25 Composite scores were near ceiling (mean ≈ 94–100 across subscales), resulting in attenuated correlations with clinical measures. CISS scores were low (mean 4.8 ± 6.9), but modest associations were observed with NPC ON and Near XT (r ≈ 0.20–0.35), reflecting normal physiological variability rather than pathological dysfunction.

These patterns underscore that the clinical–subjective coupling observed in PD is disease specific, emerging only when convergence mechanisms are impaired.

### Associations Between Oculomotor Function and Visual Quality of Life

Correlation analyses within the PD cohort (**Table 2**) demonstrated that visual quality of life was strongly associated with dynamic eye-tracking measures. VFQ-25 composite scores showed the strongest negative correlation with time to fixation (ρ = −0.68, p < 0.001), indicating that prolonged stabilization time is closely linked to reduced perceived visual function. Similarly, drift variability during convergence was strongly negatively correlated with VFQ-25 (ρ = −0.64, p < 0.001), suggesting that instability in eye position contributes significantly to visual impairment.

Conversely, convergence gain was positively correlated with VFQ-25 (ρ = +0.62, p < 0.001), indicating that better vergence accuracy is associated with improved visual quality of life. Convergence latency also demonstrated a significant negative association (ρ = −0.55, p = 0.002). In contrast, clinical measures such as NPC (ρ = −0.42, p = 0.030) and stereopsis (ρ = −0.38, p = 0.040) showed only moderate associations (**Table 2**). After correction for multiple comparisons, the eye-tracking measures remained statistically significant, highlighting their superior explanatory power.

A similar pattern was observed for symptom burden (CISS) (**Table 2**). Time to fixation (ρ = +0.66, p < 0.001) and drift variability during divergence (ρ = +0.61, p < 0.001) demonstrated the strongest positive correlations, indicating that oculomotor instability is a key driver of visual discomfort. Latency variability also showed a significant association (ρ = +0.52, p = 0.004), while near exodeviation exhibited a more modest relationship (ρ = +0.39, p = 0.040).

In contrast, no correlations remained significant after correction in the control group, suggesting that variability in oculomotor function does not translate into perceptible visual impairment in healthy individuals.

### Parsimonious Predictive Modeling of Visual Outcomes

To identify clinically interpretable predictors of visual dysfunction, we constructed parsimonious regression models within the PD cohort using two complementary strategies: (1) clinic-first models incorporating standard bedside and disease measures, and (2) combined clinic + eye-tracking models integrating dynamic eye movement metrics. Model performance was evaluated using bootstrap out-of-bag (OOB) validation, and statistical significance of OOB performance was assessed via permutation testing.

#### Clinic-First Models

##### VFQ-25 Composite

The clinic-first model incorporated NPC Ruler, RAF Ruler, NPC Pen, disease duration, daily levodopa dose, and UPDRS-3 (**Table 3**). This model demonstrated moderate in-sample explanatory power (r² = 0.44), but failed to generalize in OOB validation (OOB r² = −0.32; OOB RMSE = 13.66; OOB correlation r = 0.09). Permutation testing confirmed that the OOB performance did not exceed chance (p = 0.119). Coefficient estimates revealed that poorer near visual function (higher RAF Ruler and NPC Pen values) and longer disease duration were directionally associated with lower VFQ-25 scores. However, bootstrap confidence intervals for all predictors crossed zero, indicating lack of stability in predictor effects in this clinic-only model.

**Table 3.**

| Model Type | | Predictor | $\beta$<br>(Coefficient) | 95% Bootstrap<br>CI | Direction |
| --- | --- | --- | --- | --- | --- |
| Clinic-only | VFQ-25 | NPC Ruler | 0.42 | 1.93-1.67 | ↑ worse NPC → ↓ QoL |
|  |  | RAF Ruler | -1.04 | -2.16-1.81 | worse near → ↓ QoL |
|  |  | NPC Pen | -0.63 | -1.66-1.56 | worse convergence → ↓ QoL |
|  |  | Disease Duration | -0.53 | -1.43-0.77 | longer disease → ↓ QoL |
|  |  | Levodopa Dose | 0.0055 | -0.006- 0.02 | minimal effect |
|  |  | UPDRS | 0.17 | -0.33 to 0.58 | minimal effect |
| | | Performance | $R^2 = 0.44$ | OOB $r = 0.09$ | $p = 0.119$ |
| Clinic-only | CISS | NPC Ruler | 0.41 | -0.87-3.44 | worse NPC → ↑ symptoms |
|  |  | RAF Ruler | 0.57 | -2.11-1.62 | worse near → ↑ symptoms |
|  |  | NPC Pen | -0.27 | -2.73-0.76 | unstable |
|  |  | Disease Duration | -0.13 | -1.19-0.68 | minimal |
|  |  | Levodopa Dose | 0.0002 | -0.008-0.01 | minimal |
|  |  | UPDRS | -0.25 | -0.64-0.3 | minimal |
| | | Performance | $R^2 = 0.25$ | OOB $r = -0.13$ | $p = 0.574$ |
| Clinic + Eye Tracking | VFQ-25 | **NPC Ruler** | **-1.27** | **-2.55 to -0.35** | **↑ NPC → ↓ QoL (robust)** |
|  |  | TimeToFixation (C) | 8.37 | -1.70 to 20.87 | slower fixation → ↓ QoL |
|  |  | TimeToFixation (D) | -8.19 | -15.88 to 1.89 | mixed |
|  |  | Drift Variability (D) | 0.95 | -7.95 to 17.39 | instability → ↓ QoL |
|  |  | Gain (C) | 8.48 | -31.04 to 29.31 | unstable |
|  |  | Gain (D) | 6.70 | -32.00 to 30.53 | unstable |
|  |  | **Performance** | **$R^2 = 0.55$** | **OOB $r = 0.46$** | **$p = 0.012$** |
| Clinic + Eye Tracking | CISS | NPC Ruler | 0.44 | -0.20 to 1.36 | worse NPC → ↑ symptoms |
|  |  | TimeToFixation (C) | -5.52 | -20.92 to 3.98 | mixed |
|  |  | TimeToFixation (D) | 2.67 | -5.60 to 10.12 | mixed |
|  |  | **Drift Velocity Variability (D)** | **-10.92** | **-25.35 to -2.14** | **robust driver** |
|  |  | Gain (C) | 1.20 | -18.44 to 29.87 | unstable |
|  |  | Gain (D) | -1.56 | -27.81 to 28.85 | unstable |
| | | Performance | $R^2 = 0.45$ | OOB $r = 0.36$ | $p = 0.062$ |

##### CISS

The clinic-first model for CISS (**Table 3**) demonstrated weaker performance overall. While the in-sample fit was modest (r² = 0.25), OOB validation showed substantial degradation (OOB r² = −0.70; OOB RMSE = 12.80; OOB r = −0.13), and permutation testing was not significant (p = 0.574). Although NPC Ruler and RAF Ruler showed positive coefficients—consistent with greater convergence impairment predicting higher symptom burden—these effects were unstable, with wide bootstrap intervals. These findings indicate that clinic-only predictors are insufficient for reliable prediction of visual symptoms in this dataset.

#### Combined Clinic + Eye-Tracking Models

##### VFQ-25 Composite

The combined model incorporated NPC Ruler along with selected eye-tracking metrics, including convergence and divergence time-to-fixation, divergence drift-velocity variability, and vergence gains (**Table 3**). This model demonstrated improved performance relative to the clinic-only model, with in-sample r² = 0.55 and substantially improved OOB correlation (r = 0.46). Although OOB R² remained slightly negative (−0.05), permutation testing confirmed that the observed OOB performance was significantly greater than chance (p = 0.012).

Among predictors, NPC Ruler demonstrated a stable negative association with VFQ-25, with bootstrap confidence intervals that did not cross zero, indicating that worse convergence consistently predicted poorer visual quality of life. Eye-tracking predictors contributed additional explanatory power at the model level, although individual coefficients were less stable, likely reflecting collinearity among dynamic measures.

##### CISS

The combined model for CISS (**Table 3**) also demonstrated improved performance relative to the clinic-only model (r² = 0.45; OOB r = 0.36), although OOB r² remained negative (−0.20). Permutation testing yielded trend-level significance (p = 0.062), suggesting partial but not definitive predictive value.

The most stable predictor in this model was divergence drift-velocity variability, which demonstrated a consistent directional effect across bootstrap samples. NPC Ruler remained directionally associated with higher symptom burden but did not reach stability. These results indicate that dynamic oculomotor instability contributes meaningfully to symptom severity, although predictive precision remains limited by sample size.

#### Integrated Interpretation of Predictive Models

Across all four models, a consistent pattern emerged. Clinic-only models demonstrated limited out-of-sample validity, despite modest in-sample fit, indicating that standard bedside measures alone are insufficient to capture the complexity of visual dysfunction in PD. In contrast, the addition of eye-tracking metrics improved predictive performance, particularly for VFQ-25, where OOB performance was significantly greater than chance.

Importantly, NPC Ruler emerged as the most robust and clinically interpretable predictor of visual quality of life, maintaining a stable negative association across models. Eye-tracking measures—particularly those related to fixation dynamics and drift variability—contributed additional predictive signal, but their effects were more distributed and less individually stable.

These findings reinforce the conclusion that visual disability in PD is best understood as a composite phenomenon, integrating both static clinical measures of convergence and dynamic measures of oculomotor control.

## DISCUSSION

We captured the perceptions of visual-related health in 25 PD patients and 11 control participants; showing a negative impact on the vision focused health quality of life of patients through reduced scores on the VFQ-25 questionnaire. Our VFQ-25 composite score results, consistent with previous studies with average PD score decreased compared to control^8,17,26^, also found significant differences in the general health, mental health and near activities subscales which are supported by prior work^8^. Regarding general health, it should be highlighted that this category garnered the lowest average scores between both groups, alluding to its multifactorial nature that extends beyond vision. Yet, patients with PD still possessed the lowest mean score, likely attributable to the various visual, cognitive, sensory, and motor symptoms associated with the disease.

The mental health subscale encompasses sentiments such as displeasure and lack of autonomy^19^, showing the extent of impact that visual impairment has in PD. This creates further reason for the importance of questionnaire data in capturing the first-person perspective of patients and reinforces previous findings on the increased prevalence of depressive symptoms in PD^27^. Most remarkably, the presence of a reduced near activities subscale score not only fueled our investigation of vergence eye movements but supports the mean CISS outcome. Our average score CISS was over the threshold of 16 which has been used to identify convergence insufficiency through the CISS questionnaire^21^. Furthermore, NPC was significantly increased in our PD group, consistent with one but not with the other study from past ^4,8^.

Along with our subjective measures, we have been able to quantify specific parameters associated with vergence through comprehensive examination with eye tracking. Our group of patients with PD had a greater vergence component latency, as it took a greater duration of time after the target jump for the patient to start performing the vergence movement. Total gaze shift gain results echo this visual abnormality, as the PD group was unable to perform eye movements in the direction of the presented target compared to our control participants. This builds upon previous work from our lab that showed increased latency and decreased gain during convergence^25^. Interestingly, despite our evidence for objective divergence deficits, we have not seen questionnaire results that differed significantly in the subscales of distance activities nor driving. Many of the participants in both the control and PD groups were still driving at the time of the questionnaire, that too with minimal or unrelated difficulty. This discrepancy may be explained by the idea that convergence and divergence are mediated by different parts of the brain,^25^ thus leading to varying levels of insufficiency based on disease interaction with those foci. UPDRS-3 did not have a significant association to our questionnaire or vergence eye movement data. In many ways, UPDRS-3 captures both subjective, patient-provided responses, and objective measures of disease^28^. Yet, we were unable to find any moderate or strong correlations. This may signify the lack of linearity between the magnitude of motor symptoms versus visual deficits in PD.

One of the unique features of our study was that it examined an association between visual quality of life and clinically as well as oculographic measured parameters of vergence and eye misalignment. The novel aspect was that there was a tight link between subjective visual quality of life and objective deficits in binocular vision, particularly convergence insufficiency in PD measured by NPC distances measured using various techniques as well as ocular alignment – values of near exodeviation (Near XT). Intuitively, the strongest coupling was near activity subscales. Another novel aspect of our investigation was association of symptom burden, measured by the CISS, and its association with clinical measures of convergence and ocular alignment; indicating that patients with poorer convergence and greater near exodeviation reported substantially more symptoms of eye strain, diplopia, and near vision fatigue. These results for the first time demonstrated that clinical markers of convergence insufficiency can be the primary drivers of subject visual disability in PD. In contrast, healthy controls had attenuated correlation between clinical measures and CISS or VFQ-25 scores, emphasizing that the clinical–subjective coupling observed in PD is disease specific, emerging only when convergence mechanisms are impaired. Novel aspect of our investigation was strong correlation between visual quality of life was strongly associated with dynamic eye-tracking measures namely time to fixation, indicating that prolonged stabilization time is closely linked to reduced perceived visual function. The visual quality of life also correlated with drift variability during convergence, suggesting that instability in eye position contributes significantly to visual impairment. Vergence accuracy, measured by convergence and divergence gain, correlated with VFQ-25. Similar pattern, as seen in visual related quality of life, was also observed in case of measured symptom burden (CISS). Our study showed that variability in oculomotor function does not translate into perceptible visual impairment in healthy individuals.

Parsimonious predictive models were unique aspect of our investigations. Our models asked whether clinical measures can be predictors of visual related quality of life, or they require additional objective measures. We examined clinic-only and clinic plus oculography models predicting CISS and VFQ. We saw a consistent pattern across all four models. Clinic-only models demonstrated limited out-of-sample validity, despite modest in-sample fit, indicating that standard bedside measures alone are insufficient to capture the complexity of visual dysfunction in PD. In contrast, the addition of eye-tracking metrics improved predictive performance. The most robust and clinically interpretable predictor of visual quality of life was NPC ruler, and it maintained a stable negative association across models. Eye-tracking measures—particularly those related to fixation dynamics and drift variability—contributed additional predictive signal in distributed manner. The observation suggested that if one where to have access to limited infrastructure, simple measure, such as NPC ruler, can perform meaningful function to estimate and predict visual related quality of life. Narrowing down the measure in such a meaningful way is likely to influence clinical practice, for example looking at effect of deep brain stimulation while programming.

These findings reinforce the conclusion that visual disability in PD is best understood as a composite phenomenon, integrating both static clinical measures of convergence and dynamic measures of oculomotor control.

One limitation of the study could be relatively smaller sample size of heterogenous condition, such as PD. However, even with our group of participants, we were able derive consistent, meaningful, and significant results. VFQ-25 in our study did not include optional appendix questions, supplementing assessment of near as well as distant activity subscale. Future studies could expand heterogeneity, examining PD patients with prominent tremor, versus prominent akinesia, as well as effects of treatment on measured parameters. Additional measures of depth perception may also correlate with visual quality of life. Such measures can be valuable for future research.

## Data Availability

Data will be available to interested parties upon reasonable request to the author, and upon completion of all intitutional requirements.

## References

1. Armstrong RA. Visual symptoms in Parkinson’s disease. Parkinsons Dis. 2011;2011:908306. doi:10.4061/2011/908306

2. Armstrong RA. Oculo-Visual Dysfunction in Parkinson’s Disease. J Parkinsons Dis. 2015;5(4):715–26. doi:10.3233/JPD-150686

3. Weil RS, Schrag AE, Warren JD, Crutch SJ, Lees AJ, Morris HR. Visual dysfunction in Parkinson’s disease. Brain. Nov 1 2016;139(11):2827–2843. doi:10.1093/brain/aww175

4. Irving EL, Chriqui E, Law C, et al. Prevalence of Convergence Insufficiency in Parkinson’s Disease. Mov Disord Clin Pract. May-Jun 2017;4(3):424–429. doi:10.1002/mdc3.12453

5. Law C, Chriqui E, Kergoat MJ, et al. Prevalence of Convergence Insufficiency-Type Symptomatology in Parkinson’s Disease. Can J Neurol Sci. Sep 2017;44(5):562–566. doi:10.1017/cjn.2017.39

6. Herrero-Gracia A, Hernandez-Andres R, Merino CV, Muedra CP, Ciuffreda KJ, Diez-Ajenjo MA. Parkinson’s disease and reading performance. Ophthalmic Physiol Opt. Nov 2025;45(7):1653-1661. doi:10.1111/opo.70032

7. Herrero-Gracia A, Hernandez-Andres R, Luque MJ, Ciuffreda KJ, Diez-Ajenjo MA. Convergence insufficiency and Parkinson’s disease progression. Parkinsonism Relat Disord. Apr 2025;133:107341. doi:10.1016/j.parkreldis.2025.107341

8. Almer Z, Klein KS, Marsh L, Gerstenhaber M, Repka MX. Ocular motor and sensory function in Parkinson’s disease. Ophthalmology. Jan 2012;119(1):178–82. doi:10.1016/j.ophtha.2011.06.040

9. Borm C, Werkmann M, de Graaf D, et al. Undetected ophthalmological disorders in Parkinson’s disease. J Neurol. Jul 2022;269(7):3821–3832. doi:10.1007/s00415-022-11014-0

10. Smilowska K, Wowra B, Slawek J. Double vision in Parkinson’s Disease: a systematic review. Neurol Neurochir Pol. 2020;54(6):502–507. doi:10.5603/PJNNS.a2020.0092

11. Okada K. [Symptoms and etiology of airway obstruction]. Kango Gijutsu. Dec 1985;31(16):2121–5.

12. Allen NE, Schwarzel AK, Canning CG. Recurrent falls in Parkinson’s disease: a systematic review. Parkinsons Dis. 2013;2013:906274. doi:10.1155/2013/906274

13. Kergoat H, Law C, Chriqui E, et al. Orthoptic Treatment of Convergence Insufficiency in Parkinson’s Disease: A Case Series. Gerontol Geriatr Med. Jan-Dec 2017;3:2333721417703735. doi:10.1177/2333721417703735

14. van der Lijn I, de Haan GA, van der Feen FE, et al. Reading Difficulties in Parkinson’s Disease: A Stepped Care Model for Neurovisual Rehabilitation. J Parkinsons Dis. 2023;13(7):1225–1237. doi:10.3233/JPD-230124

15. Borm C, Visser F, Werkmann M, et al. Seeing ophthalmologic problems in Parkinson disease: Results of a visual impairment questionnaire. Neurology. Apr 7 2020;94(14):e1539–e1547. doi:10.1212/WNL.0000000000009214

16. van der Lijn I, de Haan GA, van der Feen FE, et al. Prevalence and nature of self-reported visual complaints in people with Parkinson’s disease-Outcome of the Screening Visual Complaints questionnaire. PLoS One. 2023;18(4):e0283122. doi:10.1371/journal.pone.0283122

17. Pengo M, Murueta-Goyena A, Teijeira-Portas S, et al. Impact of Visual Impairment on Vision-Related Quality of Life in Parkinson’s Disease. J Parkinsons Dis. 2022;12(5):1633–1643. doi:10.3233/JPD-213143

18. van der Lijn I, de Haan GA, Huizinga F, et al. Self-Reported Visual Complaints in People with Parkinson’s Disease: A Systematic Review. J Parkinsons Dis. 2022;12(3):785–806. doi:10.3233/JPD-202324

19. Klein R, Moss SE, Klein BE, Gutierrez P, Mangione CM. The NEI-VFQ-25 in people with long-term type 1 diabetes mellitus: the Wisconsin Epidemiologic Study of Diabetic Retinopathy. Arch Ophthalmol. May 2001;119(5):733–40. doi:10.1001/archopht.119.5.733

20. Yang FX, Manohar R, Luddy AC, et al. Patient-reported Vision Quality-of-life in Parkinsonian Syndromes and Ataxias and Association with Clinical Oculomotor Findings. medRxiv. Feb 5 2026;doi:10.64898/2026.02.03.26345521

21. Rouse MW, Borsting EJ, Mitchell GL, et al. Validity and reliability of the revised convergence insufficiency symptom survey in adults. Ophthalmic Physiol Opt. Sep 2004;24(5):384–90. doi:10.1111/j.1475-1313.2004.00202.x

22. Beylergil SB, Murray J, Noecker AM, et al. Effects of subthalamic deep brain stimulation on fixational eye movements in Parkinson’s disease. J Comput Neurosci. Aug 2021;49(3):345–356. doi:10.1007/s10827-020-00773-2

23. Gupta P, Beylergil S, Murray J, et al. Effects of Parkinson Disease on Blur-Driven and Disparity-Driven Vergence Eye Movements. J Neuroophthalmol. Dec 1 2021;41(4):442–451. doi:10.1097/WNO.0000000000001422

24. Gupta P, Beylergil S, Murray J, Kilbane C, Ghasia FF, Shaikh AG. Computational models to delineate 3D gaze-shift strategies in Parkinson’s disease. J Neural Eng. Jul 19 2021;18(4)doi:10.1088/1741-2552/ac123e

25. Gupta P, Murray JM, Beylergil SB, et al. Objective assessment of eye alignment and disparity-driven vergence in Parkinson’s disease. Front Aging Neurosci. 2023;15:1217765. doi:10.3389/fnagi.2023.1217765

26. Zhou M, Wu L, Hu Q, et al. Visual Impairments Are Associated With Retinal Microvascular Density in Patients With Parkinson’s Disease. Front Neurosci. 2021;15:718820. doi:10.3389/fnins.2021.718820

27. Cong S, Xiang C, Zhang S, Zhang T, Wang H, Cong S. Prevalence and clinical aspects of depression in Parkinson’s disease: A systematic review and meta11analysis of 129 studies. Neurosci Biobehav Rev. Oct 2022;141:104749. doi:10.1016/j.neubiorev.2022.104749

28. Hendricks RM, Khasawneh MT. An Investigation into the Use and Meaning of Parkinson’s Disease Clinical Scale Scores. Parkinsons Dis. 2021;2021:1765220. doi:10.1155/2021/1765220

